# Longitudinal automated brain volumetry vs. expert visual assessment of atrophy progression on MRI is robust but caution is advised

**DOI:** 10.1101/2024.05.21.24306349

**Authors:** Max Gebest, Christel Weiß, Chang-Gyu Cho, Lucrezia Hausner, Lutz Frölich, Alex Förster, Nandhini Santhanam, Johann Fontana, Christoph Groden, Holger Wenz, Máté E. Maros

## Abstract

Automated tools have been proposed to quantify brain volume for suspected dementia diagnoses. However, their robustness in longitudinal, real-life cohorts remains unexplored. We investigated if expert visual assessment (EVA) of atrophy progression is reflected by automated volumetric analyses (AVA) on sequential MR-imaging. We analyzed a random subset of 20 patients with two consecutive 3D T1-weighted examinations (median follow-up 4.0 years, LQ-UQ: 2.1-5.2, range: 0.2-10). Thirteen (65%) with cognitive decline, the remaining with other neuropsychiatric diseases. EVA was performed by two blinded neuroradiologists using a 3 or 5-point Likert scale for atrophy progression (scores ±0-2: no, probable and certain progression or decrease, respectively) in dementia-relevant brain regions (frontal-, parietal-, temporal lobes, hippocampi, ventricles). Differences of AVA-volumes were normalized to baseline (delta). Inter-rater agreement of EVA scores was excellent (κ=0.92). AVA-delta and EVA showed significant global associations for the right hippocampus (p=0.035), left temporal lobe (p=0.0092), ventricle volume (p=0.0091) and a weak association for the parietal lobe (p=0.067). *Post hoc* testing revealed a significant link for the left hippocampus (p=0.039). In conclusion, the associations between volumetric deltas and EVA of atrophy progression were robust for certain brain regions. However, AVA-deltas showed unexpected variance, and therefore should be used with caution in individual cases, especially when acquisition protocols vary.

## 1. Introduction

In Western countries globally, aging populations anticipate increasing socioeconomic burdens from cognitive and memory decline^1^. Structural MRI has been the main imaging modality in the early work-up of cognitive decline^2,3,4,5,6^. Recently, numerous automated brain segmentation and volumetric analysis (AVA) tools have emerged to address the shortcomings of subjective expert-based visual assessments (EVA) of brain MRIs and to assist neuro-/radiologists to quicker establish a diagnosis in the early work-up of cognitive decline^7^. Yet, their robustness in longitudinal settings and performance in comparison to neuroradiological experts in real-life follow-up scenarios remain uncertain. Thus, we investigated if expert-based brain atrophy assessment would be reflected by AVA tools in longitudinal routine clinical imaging setting.

The number of patients living with dementia is expected to triple by 2050^1^. Hence, the exploration of early biomarkers to predict a conversion to a manifested dementia in possible early stages like subjective cognitive decline^8^ and mild cognitive impairment (MCI)^9^ will be crucial in the future. Also, the National Institute on Aging and Alzheimer’s Association (NIA-AA) considers brain atrophy as a biomarker in the biological cascade of Alzheimer’s Disease^10^. Automated brain volumetry and its comparison to healthy, age-specific reference cohorts has been promised to assist clinicians in the diagnosis of dementia by highlighting the severity and distribution of brain atrophy in patients^11,12,13,14,15,16^. Such quantitative reports (Q-reports)^7^ are offered by various proprietary tools. Additionally, longitudinal volumetric approaches have shown to be able to differentiate patients with MCI from normal aging^17^, which can, therefore, potentially be used as a biomarker for diagnosing the diseases or its progression. Still, the current Canadian consensus conference on the diagnosis and treatment of dementia, advises against the routine clinical use of automated quantification software, until larger studies demonstrate their added diagnostic value^18^.

Therefore, the aim of this study was to investigate whether volumetric measurements generated by a well-established automated tool (AI-Rad Companion Brain MR version VA40 [AIRC]^19,20^, Siemens Healthineers, Forchheim, Germany) reflect brain atrophy in the same way as it is perceived during conventional visual assessment by expert neuroradiologists. To evaluate the robustness of longitudinal volumetric assessment, the qualitative and quantitative normalized volumetric differences (deltas) between baseline imaging and follow-up imaging were assessed in a random subset of twenty patients from a dementia outpatient clinic of a tertiary university provider.

## 2. Materials and Methods

### 2.1. Study population

A subset of 20 random patients was retrieved from our radiology information system (RIS) and picture archiving and communications system (PACS) who had been referred to our neuropsychiatric outpatient center (Central Institute of Mental Health, CIMH, Mannheim, Germany) for MR-imaging and follow-up examination between 06/2022 and 08/2022. Patients with at least two time points (TP) with imaging were included. A thin-slice (1mm), three-dimensional (3D) T1-weighted gradient echo sequence (magnetization prepared rapid acquisition with gradient echoes, MPRAGE) was part of the imaging protocol.

The study was approved by the Medical Ethics Commission II of Medical Faculty Mannheim, University of Heidelberg (approval nr.: 2017-825R-MA and 2017-828R-MA). The need for informed consent was waived because of the retrospective nature of the study. All procedures were in accordance with the ethical standards of the institutional and national research committee and with the 1964 Helsinki Declaration and its later amendments or comparable ethical standards.

### 2.2. Imaging protocol

For all patients, a set of conventional sequences was available, which included 3 mm transversal fluid attenuated inversion recovery (FLAIR), diffusion-weighted imaging (DWI) with apparent diffusion coefficient (ADC), T2-weighted turbo spin-echo, T2* or susceptibility weighted imaging (SWI) and an isotropic 3D T1 MPRAGE (voxel size: 1 x 1 x 1 mm^3^). MRI scans were performed on three different MRI-scanners, including 3T MAGNETOM Trio, MAGNETOM Prisma and Biograph mMR PET-MR system (all from Siemens Healthineers AG, Erlangen, Germany) with slightly varying protocol parameters for the 3D T1 MPRAGE-sequence. Relevant parameters for the automated volumetric software tool are summarized in **Supplementary** Error! Reference source not found..

### 2.3. Expert-based visual assessment (EVA)

Human expert-based visual assessment was set as the gold standard. Two independent and blinded radiologists with advanced and intermediate experience (H.W. 13 years; M.G. 5 years) performed visual assessment. Images were conventionally compared side-by-side using open-source DICOM viewer (Horos®, v.4.0.0, https://horosproject.org) in the default clinical work environment. Qualitative structured evaluation was performed as suggested^21^ using standardized slices of assessment of the 3D T1 MPRAGE for hippocampus^22^, parietal^23^, frontal and temporal^24^ lobes. Consequently, seven different anatomical brain regions, relevant for the diagnosis of dementia, were rated for atrophy progression, including: frontal lobe, parietal lobe (both sides considered as one region), supratentorial ventricles (lateral ventricles and 3^rd^ ventricle, considered as a single region), right and left temporal lobes, as well as right and left hippocampus.

A 3-point Likert scale was employed to assess longitudinal parenchymal atrophy between two MRI surveys: score 0 indicating no change in atrophy over time, score 1 indicating probable progressive atrophy and score 2 indicating certain progressive atrophy. For the ventricles, the scale was expanded with two additional scores: score 0 representing no change, score 1 for probable enlargement, score 2 for certain enlargement, score -1 for probable narrowing, and score -2 for certain narrowing of ventricles between measurements. Every region was scored individually. Discrepant scoring values were reviewed and discussed by both readers until consensus was reached.

### 2.4. Automated brain volumetric analyses (AVA)

For automated brain segmentation and volumetry, the AIRC-tool (v. VA40 Siemens Healthineers AG, Forchheim, Germany) was used due to the streamlined integration opportunity from bottom-up (MR device and end-analysis). The brain segmentation and volumetry were derived from 3D T1-weighted MPRAGE sequences according to our institutional protocol, which was not fully compliant to Alzheimer’s Disease Neuroimaging Initiative (ADNI) recommendations, as required by the FDA-approved proprietary software tool AIRC. It analyzed 51 different brain regions in both hemispheres and white matter lesions resulting in a total of 52 (sub)regions. The volumetric values were internally compared to the built-in age- and sex-matched reference database of the AIRC tool consisting of 303 healthy individuals. Segmentation and volumetric results were presented in a fused image projected on top of the input 3D T1 MPRAGE image as i) a deviation map showing color-coded z-scores (10th–90th percentiles) estimated from the included reference cohort and ii) a color-coded map that showed the segmentation results of anatomical regions. Variations in individual head sizes were corrected by normalization to the total intracranial volume (TIV) of the respective patient. The volumetric results were displayed as the absolute and TIV-normalized values.

### 2.5. Comparison and matching of EVA and AVA

The AIRC-tool provides distinct volumetric measurements for 52 anatomical subregions including laterality (left and right side) and the gray and white matter. Thus, these AIRC-subregions had to be grouped and matched to the seven brain regions (frontal, parietal, left/right temporal lobe, left/right hippocampus and ventricle volume), which were evaluated during EVA-scoring (**Supplementary Error! Reference source not found.**).

To compare the results of the visual assessment and the results of the quantitative assessment, we calculated the normalized difference (delta) of the absolute volume for each of the seven brain regions between the two imaging timepoints (TP) and normalized it to the region’s baseline (TP1)^25^.

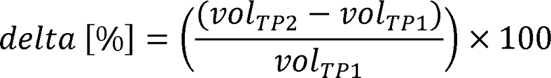

A positive delta in percent indicated an increase in the volume of the respective anatomical region, while a negative delta in percent suggested a decrease in normalized volume.

### 2.6. Statistical analyses

Non-normally distributed variables were described with their medians, lower and upper quartile (LQ-UQ) while proportions were shown for categorical variables. Statistical analyses were conceptualized by (M.M) and performed by (C.W.) using SAS for Windows (v.9.4, Statistical Analysis System, SAS Institute, Cary, North Caroline, USA). All statistics were non-parametric according to group sizes. To assess statistical associations, first, global- (Kruskal-Wallis, KW)^26^ then *post hoc* tests using paired Wilcoxon-signed rank test (WSRT) were performed. Due to limited group sizes of certain anatomical regions, EVA scores of 1 and 2 were combined for subsequent sensitivity analyses. Distributions of follow-up times between groups were compared using unpaired two-samples Wilcoxon-Mann-Whitney (WMW) tests. Significance thresholds were adjusted for multiple testing using either the Bonferroni- or Holm-method^27,28,29^ (n_test_=12). Adjusted p*<0.0042(=0.05/12) were considered significant. Inter-rater agreement was calculated using weighted Cohen’s kappa (κ) statistics for two ordinal variables. Figures were created (M.G.) using GraphPad Prism (v. 10.0.3, GraphPad Software LLC, Boston, USA).

## 3. Results

### 3.1. Study cohort

In total, a random subset of 20 patients (n=14 female, 70%; **Table 1**) with median age of 66.1 years (LQ-UQ: 58.1-73.9, range: 36.4-90.2 yrs) was included from the memory and neuropsychiatric outpatient clinic of the Central Institute of Mental Health, Mannheim and Dept. of Neuroradiology, Medical Faculty Mannheim, University of Heidelberg, Mannheim, Germany. Patients underwent memory-/cognitive decline work-up (n=13; 65%) or a diagnostic routine evaluation for neuropsychiatric diseases (n=7; 35%) such as schizophrenia and depression. All patients received at least two consecutive MR-imaging studies. Overall, the 40 MRIs were all performed on 3T MRI machines (MAGNETOM Trio n=14; MAGNETOM Prisma n=14; Biograph mMR PET-MR system, n=12, all Siemens Healthineers AG, Erlangen, Germany). Only 5/20 patients were scanned on the same MRI-machine for both MRI-surveys (n=2 Prisma/Prisma; n=2 Biograph/Biograph; n=1 Trio/Trio), while 15 patients were scanned on two different MRI machines (n=3 Prism/Biograph; n=7 Trio/Prisma; n=5 Trio/Biograph) and thus different 3D T1 protocol parameters were used (**Supplementary Table S1**).

**Table 1.** Summary of patient demographics and clinical characteristics.

|  |  | **Clinical diagnoses** |  |
| --- | --- | --- | --- |
|  | **Whole sample** | **MCI** | **Other neuropsychiatric diseases** |
| **Sample size (%)** | 20 | 13 (65) | 7 (35) |
| **Sex (female; %)** | 14 (70) | 7 (53.8) | 7 (100) |
| **Age median** | 66.1 | 69.0 | 63.1 |
| **(LQ-UQ; range [yrs])** | (58.1-73.9; 36.4-90.2) | (59.3-75.2; 54.9-90.2) | (54.1-72.2; 36.4-80.7) |
| **Median follow-up** | 4.0 | 4.1 | 3.3 |
| **(LQ-UQ; range [yrs])** | (2.1-5.2; 0.2-10) | (2.2-5.3; 0.2-10) | (1.9-4.0; 0.8-5.6) |
| **N scanned on same scanner (%)** | 5 (25) | 3 (60) | 2 (40) |

Population characteristics for the dataset of 20 random patients, who had been referred to our neuropsychiatric outpatient center at the Central Institute of Mental Health CIMH, Mannheim, Germany for imaging follow-up between 06/2022 and 08/2022. MCI: mild cognitive impairment.

All AVA-segmentation results presented in a fused image projected on top of the input 3D T1 MPRAGE image were inspected if available. Segmentation irregularities (**Supplementary Figure S1**) were found frequently and continuously distributed in terms of their severity. Note that the software did not allow manual edits to the segmentation results as results were directly sent to the PACS.

### 3.2. Imaging follow-up time

Median imaging follow-up time for the whole sample was 4.0 years (LQ-UQ: 2.1-5.2 yrs, range: 0.2-10 yrs). Median imaging follow-up time for patients with cognitive decline was 4.1 (LQ-UQ: 2.2-5.3yrs, range: 0.2-10 yrs) and with 3.3 years slightly shorter (p_WMW_=0.35) for patients with other neuropsychiatric diseases (LQ-UQ: 1.9-4.0 yrs, range: 0.8-5.6 yrs). It is noteworthy that there was one patient with imaging follow-up under one year, conducted on the same MRI-device, in the MCI group (0.9 yrs) and one in the neuropsychiatric diseases group (0.8 yrs), respectively. There was no relevant (p_WMW_=0.55) difference between the imaging follow-up time of male (median 4.2 yrs; LQ-UQ: 4.1-4.9 yrs, range: 0.2-7.8 yrs) or female (median 3.3 yrs, LQ-UQ: 1.9-5.2 yrs, range: 0.8-10 yrs) patients.

### 3.3. Inter-rater agreement of expert visual assessment (EVA) scores

Average weighted κ over all parenchymal regions showed a high inter-rater agreement (κ=0.92). This included the frontal lobe (κ=1.0), parietal lobe (κ=0.83), right temporal lobe (κ=0.85), right hippocampus (κ=0.91), left temporal lobe (κ=0.93) and left hippocampus (κ=0.92), as well as ventricles (κ=0.92). For the ventricles we had to include score -1 and score -2 to the Likert scale, since one patient clearly showed a narrowing of the ventricles over time without any structural cause like a tumor or iatrogenic intervention like ventricle drainage.

### 3.4. Group comparisons between expert visual assessment (EVA) scores and automated brain volumetric analyses (AVA) delta

Results of EVA scoring and delta of AVA in percentage are displayed in **Table 2**. There was a large spread of deltas between the two MRI surveys for each of the seven regions that are relevant for the diagnosis of dementia.

**Table 2.** Summary of the investigated brain regions comparing expert visual assessment (EVA) scores and AI-based brain volumetric analyses (AVA).

| Brain regions | EVA |  | AVA-delta* | p<sub>KW</sub> |
| --- | --- | --- | --- | --- |
|  | score | n (%) | mean, (min.; max.) |  |
| frontal lobe | 0 | 12 (60%) | +5.3 (-12.7; +10.6) | 0.63 |
|  | 1 | 3 (15%) | -2.6 (-6.4; +2.3) |  |
|  | 2 | 5 (25%) | +7.8 (-8.1; +13.5) |  |
| parietal lobe | 0 | 16 (80%) | +1.7 (-4.6; +18.6) | 0.067 |
|  | 1 | 2 (10%) | +4.7 (-4.9; +14.3) |  |
|  | 2 | 2 (10%) | -4.9 (-8.2; -1.5) |  |
| left temporal lobe | 0 | 13 (65%) | -0.8 (-9.5; +12.6) | **0.0092** |
|  | 1 | 3 (15%) | -8.3 (-12.9; -5.6) |  |
|  | 2 | 4 (20%) | -9.6 (-15.1; -5.3) |  |
| right temporal lobe | 0 | 14 (70%) | -0.3 (-11.9; +12.0) | 0.11 |
|  | 1 | 3 (15%) | -2.6 (-3.9; -0.7) |  |
|  | 2 | 3 (15%) | -7.2 (-12.1; -4.6) |  |
| left hippocampus | 0 | 15 (75%) | -0.6 (-18.5; +21.7) | 0.12 |
|  | 1 | 2 (10%) | -10.7 (-10.8; -10.5) |  |
|  | 2 | 3 (15%) | -12.1 (-24.1; -5.9) |  |
| right hippocampus | 0 | 15 (75%) | +1.0 (-12.0; +27.8) | **0.035** |
|  | 1 | 3 (15%) | -9.5 (-11.5; -8.1) |  |
|  | 2 | 2 (10%) | -10.8 (-18.5; -3.0) |  |
| ventricles | 0 | 10 (50%) | +10.2 (-7.1; 24.4) | **0.0091** |
|  | 1 | 3 (15%) | 16.7 (-3.1; +42.8) |  |
|  | 2 | 6 (30%) | 37.7 (+26.0; +66.0) |  |
|  | -1 | 0 (0%) | --- |  |
|  | -2 | 1 (5%) | -28.6 (-28.6; -28.6) |  |

AVA: AI-based brain volumetric analyses; \*volumetric differences (deltas, %) were generated using the AI Rad Companion, Brain MR Tool (v. VA2x, Siemens Healthineers GmbH, Erlangen, Germany). EVA: expert visual assessment; p<sub>KW</sub> were calculated using the Kruskal Wallis test; **bold** indicates significance at p < 0.05.

For each of the seven key anatomical regions, we calculated a global non-parametric test score to correlate EVA-scores with normalized region-specific AVA deltas. The link between EVA and AVA for the frontal lobe (p_KW_=0.63, **Figure 1a**) could not be statistically verified. We found weak significance (p_KW_=0.067, **Figure 1b**) for the parietal lobe. The right temporal lobe (p_KW_=0.11, **Figure 1c**) did not show significant association, while there were strong associations for the for the left temporal lobe (p_KW_=0.0092, **Figure 1d**) and the right hippocampus (p_KW_=0.035, **Figure 1e**). There was no association for the left hippocampus (p_KW_=0.12, **Figure 1f**). Lastly, we found strong association for the ventricle system (p_KW_=0.0091, **Figure 1g**) between EVA and AVA. The delta differences between ventricle size (p=0.0002) for EVA score 0 and 2 stayed significant after Bonferroni and/or Holm-correction (p*=0.0042).

**Figure 1.**
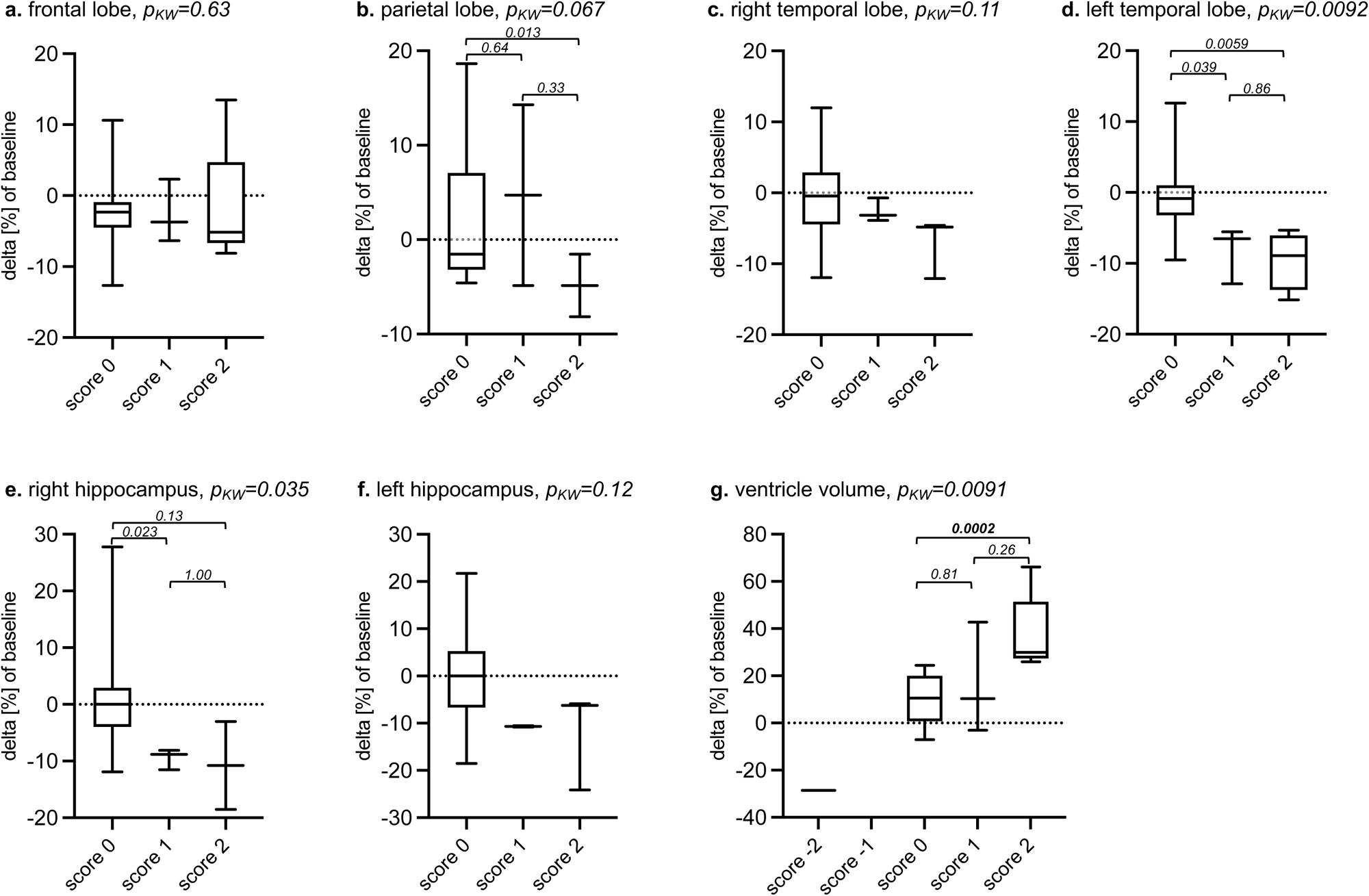
Combination figure of expert visual assessment (EVA) scoring of atrophy between baseline and follow-up vs. automated volumetric analysis (AVA). The x-axes depict EVA grouped as score 0: no perceptible atrophy; score 1: probable progressive atrophy; score 2: certain progressive atrophy for the (a) frontal-, (b) parietal-, (c) right temporal-, (d) left temporal lobe, the (e) right and (f) left hippocampus while for the (g) ventricles score 0: represented no change, score 1: probable enlargement, score 2: certain enlargement, score -1: probable narrowing, and score -2: certain narrowing. Y-axes depict normalized difference (delta [%]) of AVA measurements between baseline and follow-up imaging. Kruskal-Wallis test (KW) revealed significant global associations between EVA and AVA-deltas for the left temporal lobe (d, p_KW_ =0.0092), the right hippocampus (e, p_KW_ =0.035), and for the ventricle volume (g, p_KW_ =0.0091). Pairwise comparisons (dashes) between EVA scoring levels 0 and 1 were significant for left temporal lobe (d, p=0.039) and right hippocampus (e, p=0.023). Pairwise comparisons between EVA scoring levels 0 and 2 were significant for the parietal lobe (b, p=0.013), left temporal lobe (d, p=0.0059) and ventricle volume (g, p=0.0002). Bold indicates significance at p* after Bonferroni-Holm-corrections. Pairwise comparisons were not performed for the (a) frontal lobe, (b) parietal lobe, the (c) right temporal lobe and the (f) left hippocampus since the KW test did not reach significance.

As sensitivity analyses, when combining EVA suspected and certain progression (scores 1 and 2) vs. no atrophy (score 0) progression, *post hoc* tests revealed a link between the left hippocampus (p_WSRT_=0.039), right hippocampus (p_WSRT_=0.0072), left temporal lobe (p_WSRT_=0.0012) and ventricle volume (p_WSRT_=0.0076). The parietal lobes (p_WSRT_=0.064) showed a weak trend towards a potential association while the right temporal- (p_WSRT_=0.13) and the frontal lobes (p_WSRT_=0.43) missed significance.

### 3.5. Robustness of longitudinal automated brain volumetric analysis in individual cases

In a subset of cases in which we could not detect atrophy over time during expert visual assessment (score 0), there was still a wide range of delta on AVA measurement. For example, in the right hippocampus we found a range of +27.8% to -12%. For variance of AVA results in all regions see **Table 2**. Summary of the investigated brain regions comparing expert visual assessment (EVA) scores and AI-based brain volumetric analyses (AVA).

On the one hand one patient out of the MCI group was scanned twice within 11 months on the Biograph MRI scanner. In the EVA, no atrophy progression was found, resulting in EVA scores of 0 across all regions. However, AVA revealed notable deltas in parenchymal regions: frontal +13.5%, parietal +10.8%, right temporal +12%, left temporal +12.6%, right hippocampus +27.8%, left hippocampus +21.7%. On the other hand, one patient out of the group with other neuropsychiatric diseases who was scanned twice within 10 months on the Prisma MRI scanner, exhibited smaller, more plausible deltas in most parenchymal regions, except for the left hippocampus (frontal -1.1%, parietal -0.84%, right temporal -1.0%, left temporal -0.84%, right hippocampus 0%, left hippocampus +8.6%).

## 4. Discussion

In this real-life explorative study, we investigated if expert visual assessment of longitudinal brain atrophy would be reflected by quantitative volumetric measurements produced by the AIRC-tool, an FDA-cleared, commercially available volumetric software application. Our study described three key findings regarding EVA-based atrophy progression assessment vs. longitudinal AVA: (1) we found a strong association between atrophy assessment by human experts and the normalized deltas of automated volumetric measurements, particularly for the right hippocampus, left temporal lobe and the ventricle system; (2) automated volumetric deltas between imaging timepoints showed high variance and were often not reflected by EVA-scores; although (3) EVAs by human experts showed strong inter-rater agreement. Hence, our findings underline the importance of carefully evaluating results of volumetric automated tools, if used for longitudinal assessment with varying acquisition protocols.

The significant association between EVA and AVA of the right hippocampus might be attributed to the highly standardized visual assessment procedure of the hippocampi, which showed high inter- and intra-rater reliability in a meta-analysis^30^. Our results are also in line with results by Velickaite *et al.,* where they showed significant association between the conventional brain atrophy score for medial temporal atrophy (MTA) and volumetric assessment in a cohort of 201 cases with well-preserved cognitive function at the age of 75 and 80 years^31^. Rau *et al.* also emphasized the robustness and clinical reliability of the MTA score^32^. Similarly, we found that besides the hippocampus, the anatomically closely associated left temporal lobe was significantly linked between human readers and the automated tool. Also, ventricle volume showed a strong association between EVA and AVA. This is relevant for patients with normal pressure hydrocephalus (NPH), a condition characterized by the accumulation of excess cerebrospinal fluid, leading to ventricular enlargement. NPH is one of the few treatable forms of dementia. Furthermore, Rau *et al.* showed that a machine learning algorithm could reliably detect NPH-patterns in 3D T1 MPRAGE images^33^.

Visual assessment was found to be less sensitive than volumetric analysis using open-source or commercial tools^31,34^. We found a weak correlation between EVA and AVA for the parietal and no association for the frontal lobe. This might be because, in our cohort, the median age was lower (66 years) than in other studies^31^. Also, median imaging follow-up time was 4 years in the whole cohort, and slightly longer in the MCI-compared to the neuropsychiatric disease group. This might have affected expert performance as longer follow-up periods increase the likelihood of detecting atrophy progression by EVA, as previously described for MTA scoring in the hippocampus^34^.

The lack of association between EVA and AVA in certain regions might be attributed to inherent shortcomings of visual assessment, and might be particularly insensitive for the frontal and parietal lobes^31^. In contrast, inter-rater agreement was highest for the frontal lobe in our cohort.

When comparing AVA-results generated by the AIRC tool to measurements from the open-source FreeSurfer tool in 45 patients with *de novo* symptoms of memory decline, Rahmani *et al.* found excellent-to-good intraclass correlation consistency between the two tools in measured absolute volumes. They concluded that the AIRC-tool reliably detects atrophy in cortical and subcortical regions relevant for diagnosing dementia^35^.

The metric used in this study, the percentage delta of the absolute volumes by the AVA, may produce higher numeric values for smaller brain regions due to its inverse proportionality to the baseline volume. Whitwell *et al.* suggested normalizing AVA measurements to total intracranial volume (TIV) for both cross-sectional and longitudinal studies to account for inter- and intraindividual image differences^36^. They found that TIV normalization reduced inter-image differences caused by voxel-scaling variations from 1.3% to 0.5% (p=0.002). Although this reduction was statistically significant, the degree of variation was quite small. Thus, it should not have introduced substantial error into our results. Moreover, the right hippocampus, despite being a small structure, emerged as one of the most robust anatomical region in terms of consistency between EVA and AVA. Furthermore, its variance for each EVA scoring level (**Table 2**. Summary of the investigated brain regions comparing expert visual assessment (EVA) scores and AI-based brain volumetric analyses (AVA).) stayed comparable to large lobar structures and was smaller than for the entire ventricle system.

Zaki *et al.* compared two different AI algorithms for normative brain volumetry in 60 MCI patients and 20 controls. They found that different algorithms can have distinct effects that impact clinical interpretation, when used in isolation. Furthermore, they concluded that these AI-tools are not interchangeable during follow-up and need internal evaluation before adoption^37^. Recently, many of these commercial AI-tools despite being CE/FDA approved, lacked proper clinical validation, especially for in-use evaluation^7^. Pemberton *et al.* showed that the use of Q-reports alongside visual assessment improved sensitivity, accuracy, and inter-rater agreement for the detection of volume loss^13^. They found that Q-reports were most useful for consultant-level radiologists, implying that more experience is required to benefit from information provided by quantitative analyses. Similarly, our findings showed that longitudinal AVA-based assessment may exhibit unexpected variability and should not be evaluated in isolation, in particular, when different acquisition protocols are being used. The results must be carefully inspected regarding technical limitations on a case-by-case basis. In a subset of cases in which EVA did not show atrophy progression in the frontal lobe and hippocampi, we found substantial variations with improbable volume gains and losses. This variation was >±10% for the frontal lobe and >±20% for the left hippocampus during a median follow-up of four years. In our experience with AIRC, the frontal and less often the parietal- and the temporal lobes were prone to segmentation instability of different severity. Some of these inconsistencies might be caused by skull-stripping, for which various approaches have been described and implemented^38,39^. Consequently, parts of the brain may not be recognized as parenchyma and are not incorporated into the volumetric measurements (i.e., “minus-variant”, **Supplementary Figure S1**). In other cases, parts of the skull, dura, falx cerebri and venous sinuses were additionally incorporated leading to a volumetric surplus (i.e., “plus-variant”, **Supplementary Figure S1**). To note, however, that the most severe cases were almost exclusively “minus”-variants, particularly, in the frontal lobe. Therefore, to rule out segmentation errors, quantitative analytics results should be inspected thoroughly and discarded if “minus” or “plus”-variants are observed. This leads to the question what margin of error can be considered acceptable, when published annual atrophy rates based on normative populational data are around and under 1%. In their meta-analysis, Fraser *et al.* showed a mean hippocampal atrophy rate as low as 0.85% per year in a sample of n=3422^40^. Sluimer *et al.* published annualized whole brain atrophy rates of -0.5% for healthy controls and -1.2% for patients with MCI^41^. If employed in longitudinal assessment the volumetric results from AIRC grossly surpass anticipated atrophy rates for the follow-up duration, or even suggest a volumetric gain, caution is advised during interpretation.

This high variance might also be explained by methodological and technical factors that can impact automated voxel-based morphometry. These factors have been previously described in detail including field strength, image resolution, acquisition sequence and image quality^42^. A study by Haller *et al.* showed that basic sequence parameters systematically bias volume estimation^36^. Huppertz *et al.* investigated intra- and inter-scanner variability of brain volumetry multiple scans of a young, healthy volunteer in six different scanners^43^. Reproducibility was best when the same sequence-protocol was performed on the same MRI-scanner. In our study, the use of different sequence-preferences and different MRI-machines may have impacted the volumetric results of the AIRC-tool. Although all three scanners were manufactured by the same vendor, even scanning on the same MRI-machine for baseline and follow-up did not fully rule out implausible AVA results. Scanning twice on Prisma produced more plausible results than scanning twice on Biograph. Notably, in patients with short follow-up time, in whom no severe atrophy progression was expected and accordingly none was detected during EVA, we found considerable variance of intraindividual regional volumetric changes. This might be counteracted by establishing reference atrophy ranges for image pairs using regularization techniques and Bayesian analytics, particularly for short term follow-ups of < 1 year^44^.

Hydration status is also known to influence brain volumetry results^45, 46, 47^, which might need further investigation in the context of AIRC robustness.

The present study has certain limitations, as it is a retrospective, explorative study of small size. Thus, it might be underpowered to confirm associations for all regions that are relevant for the diagnosis of dementia. However, it represents a random cohort of the daily practice. Similarly, all the imaging studies were performed on MR-scanners from the same vendor (Siemens Healthineers), but inevitably additional variance was introduced since follow-up images were acquired on various MRI scanners, as expected in the daily clinical praxis. Additionally, scan settings were not ADNI-conform, which potentially further limits the accuracy of the investigated proprietary AIRC tool. To note, however, that non-ADNI-conform settings represent the reality of many imaging centers and praxes. Lastly, the metric of percentage delta of AVA inherently yields higher numeric differences in smaller brain regions. However, the AVA-delta variances of the hippocampus were comparable to or even smaller than those of lobar structures or the entire ventricle system, regardless of the EVA-scoring level.

In conclusion, we found substantial and robust associations between human expert visual assessment and atrophy progression measured by the AIRC-tool for the right hippocampus, left temporal lobe and the ventricle system. Despite EVAs showing strong inter-rater agreement between human readers, like for the frontal lobe, normalized automated volumetric differences (deltas) between imaging timepoints often did not align with EVA-scores and exhibited substantial variance, even implying volume gain. Hence, our findings highlight the importance of carefully evaluating volumetric results of AIRC, when used for longitudinal assessment, especially when volumetric deltas exceed expectable atrophy rates for the duration of the follow-up. Caution is advised and results should be critically reviewed, especially when acquisition protocols vary across scans.

## 5. Data availability statement

The dataset used in this study is available from the corresponding author upon reasonable request.

## Data Availability

All data produced in the present study are available upon reasonable request to the authors.

## Acknowledgements

We are grateful to Dr. Bénédicte Maréchal (R&D Integrated Decision Support Team Manager, Advanced Clinical Imaging Technology, Siemens Healthineers) and Dr. Tobias Kober (Director Innovation Hub Switzerland, Siemens Healthineers) for their valuable comments and helpful insights.

## Authors’ contributions

M.G. and M.E.M conceptualized the study. M.G. collected imaging data. L.H., L.F. provided clinical data. M.G. and H.W. performed visual image analysis. M.G., C.GC., H.W., M.E.M performed volumetric data extraction. C.W., M.G., M.E.M. performed statistical analyses. M.G., C.W., N.S., H.W., M.E.M. analyzed and interpreted the data. M.G. created figures. L.H., L.F., A.F., J.F., C.G. advised clinical aspects of the study. M.G., H.W., and M.E.M wrote the manuscript. H.W. and M.E.M. supervised the study. All authors critically reviewed the manuscript and approved the final version.

## Figure legends

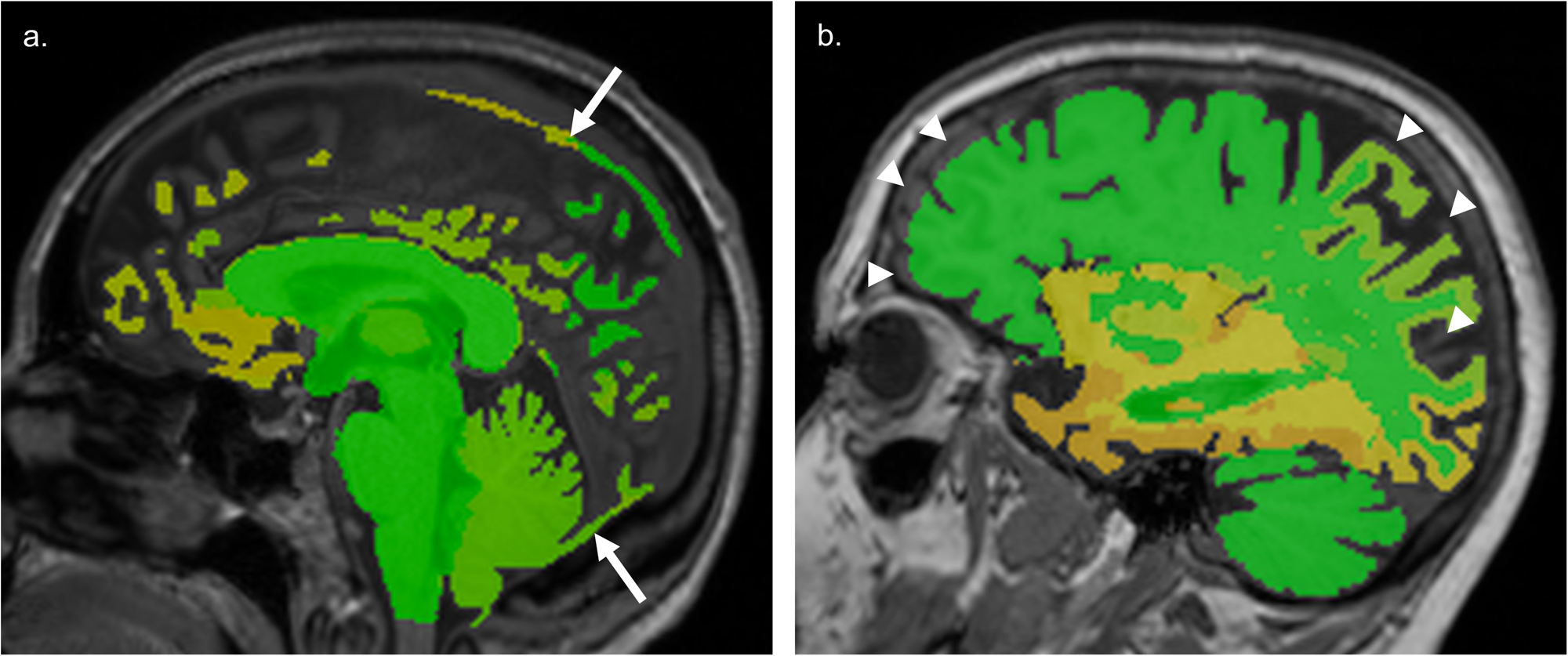

