## Supplementary materials for "Longitudinal automated brain volumetry vs. expert visual assessment of atrophy progression on MRI is robust but caution is advised"

| **Supplementary Table S1.** Device settings for 3D T1 MPRAGE on different MRI-scanners. | | | | | |
| --- | --- | --- | --- | --- | --- |
| **MRI scanner*** | **Inversion time (ms)** | **Flip angle (°)** | **Repetition time (ms)** | **Pixel bandwidth** | **Number of imaging per scanner** |
| 3T Biograph mMR PET-MR system | 1040 | 9 | 1830 | 180 | 12 |
| 3T MAGNETOM Prisma | 900 | 8 | 1800 | 160 | 14 |
| 3T MAGNETOM Trio | 900 | 9 | 1900 | 199 | 14 |
| *Devices are from Siemens Healthineers/Healthcare GmbH, Erlangen Germany. Abbreviations: T, Tesla, magnetic field strength. | | | | | |

| **Supplementary Table S2**: Summary table of anatomical regions provided by the AIRC-tool that were grouped for comparison with expert visual assessment-score. | | | | | | |
| --- | --- | --- | --- | --- | --- | --- |
| **frontal lobe** | **parietal lobe** | **left temporal lobe** | **right temporal lobe** | **left hippo-campus** | **right hippo-campus** | **ventricles** |
| frontal GM right | parietal GM right | temporal GM left | temporal GM right t | hippo-campus left | hippo-campus right | lateral ventricle left |
| frontal GM left | parietal GM left | temporal WM left | temporal WM right |  |  | lateral ventricle right |
| frontal WM right | parietal WM right |  |  |  |  | 3rd ventricle |
| frontal WM left | parietal WM left |  |  |  |  |  |
| Abbreviations: GM: gray matter, WM: white matter | | | | | | |

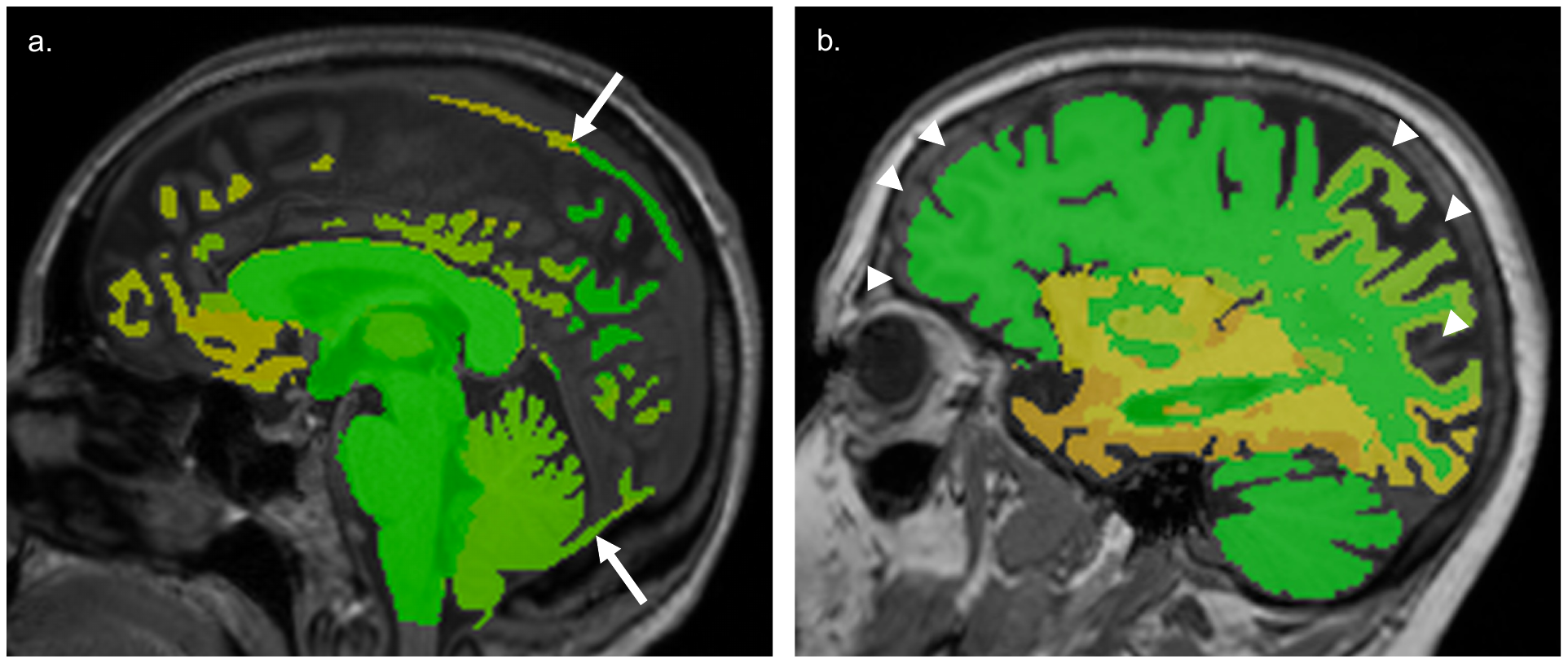

**Supplementary Figure S1**. Examples of segmentation irregularities. Fused image of 3D T1 MPRAGE and color-coded deviation map (z-scores) provided by the AIRC tool. a. Arrows pointing to “plus-variant”, where the AIRC-tool included parts of the falx cerebri and venous sinuses. b. Arrowheads pointing to “minus-variant”, where the tool missed parts of the frontal and parietooccipital lobes.
